# Time to resolution of Long COVID symptoms and the associated factors among the people infected with SARS-CoV-2 during the 3^rd^ wave of COVID-19 in Zambia: A prospective quantitative study

**DOI:** 10.64898/2026.10.28.26366684

**Authors:** Sialubanje Cephas, Phyllis I. Sumbwa, Kunka Evaristo, Nawa Mukumbuta, Mukonka Victor

## Abstract

This study was conducted over 24 months (September 2021-Agust, 2023) to determine the time to resolution of Long COVID symptoms and the associated factors among people infected with SARS-CoV-2 during the 3rd wave (June - August, 2021) of COVID-19 in Zambia. Participants comprised a cohort of 303 people randomly sampled from 7 healthcare facilities from Lusaka and Southern provinces. Demographic and clinical data were collected at baseline and at a monthly interval from participants’ clinical records and interview. Descriptive analysis was conducted to summarise respondent demographic and clinical characteristics. Survival analysis (Kaplan Meier model) was conducted to determine the median time to symptom resolution, generate survival curves and the risk table. A Poisson model was fitted to compute the crude incident rate of recovery. Cox-Proportional Hazards regression analysis was conducted to determine the univariate and multivariate association between the covariates and the time to resolution of symptoms. Crude and adjusted hazard ratios, 95% confidence intervals and p-values were computed. A total of 236 events occurred during the follow- up period; the crude incidence rate was 1, 575.6 events per 1000 person years (95%CI: 1,363.2 - 1,784.4). The median time from diagnosis of Long COVID symptoms to resolution was 6 (IQR: 5-8) months. Vaccination status was significantly associated with the time to resolution of symptoms (aHR= 1.71, 95% CI: 1.02 - 2.86, p=0.04) after adjusting for age, number of children, occupation, pre-existing medical conditions, number of vaccine doses and hospital admission. Other variables did not show statistically significant association with the time to resolution of symptoms (p>0.05). These findings suggest that vaccination is associated with a shorter time to resolution of Long COVID symptoms. Netherless, causation can not be ascertained due to the observational design of the study. A study with an experimental design is needed to establish the causal relationship.

## Introduction

Globally, as of 28 August 2022, there were over 598 million confirmed cases of COVID-19 and over 6.4 million deaths [1]. In Zambia, a total of 325, 248 confirmed cases and 4,003 deaths were reported by this time [2]. Although high case fatality rates were reported from developed countries during successive waves [3], most infections in Zambia were mild with an overall case fatality rate of 1.23%. However, the third wave (June to August 2021), reported more cases than the previous waves, with a cumulative total of 206,051 cases, 3,596 deaths and a case fatality (CFR) of 1.75%. The severe cases and CFR were attributed to the delta variant [4].

Although most people typically recovered from COVID-19 after 2 to 6 weeks [5], studies and reviews [6,7] have demonstrated that some symptoms may recur or persist for weeks or months following initial recovery among both the healthy people and those with mild disease. Some medical complications may have lasting health effects on the survivors; some patients may not regain their full health [8]. These lasting health effects have been collectively termed Long COVID or Post COVID conditions–defined by the World Health Organisation (WHO) as “symptoms appearing 3 or more months post-infection with SARS-CoV-2 infection, lasting for 2 or more months and not explained by an alternative diagnosis [9]. According to WHO [9], around 10–20% of people infected with SARS-CoV-2 may go on to develop symptoms that can be diagnosed; approximately 10-15% of cases progress to severe disease; 5% become critically ill with severe clinical manifestations and complications ranging from shortness of breath and general breathing difficulties to organ failure and/or death. Respiratory and cardiac complications have been reported to be the most common complications and leading causes of death, especially among the elderly and those with co-morbidities [10].

Although elderly patients and those with co-morbidities have a high probability of developing complications [11,12], a CDC report [13] indicates cases of healthy adults and young people who developed Long COVID-related complications. Reports show that many people may have experienced it during the first two years of the pandemic (2020/21); however, the exact number of those experiencing or living with the condition is uncertain due to health system challenges in low-and-middle income countries (LMICs) [14]. Consequently, many people who suffered/ and or are living with the Long COVID condition(s) may go unreported or undiagnosed [14,15]. A study conducted in Zambia reported a Long COVID prevalence of 26% at the end of a one year follow up period.

Due to scarcity of Long COVID data from prospective African cohorts to highlight novelty, much is still unknown with regard to: a) which and what proportion of patients will have some symptoms persist, recur or resolve; b) how long the symptoms persist or take to resolve; c) the long-term effects of the disease, and d) the associated factors. Most published research were conducted in developed countries; others focused on the biology of the virus itself, prevention and control measures, clinical manifestations of the disease and other related aspects; little attention has been paid to the long-term effects of the disease. In view of these important knowledge gaps, a prospective study was conducted among a selected cohort of COVID-19 survivors with mild, or no symptoms during the 3^rd^ wave of COVID-19 in Zambia.

The aim of the study was to determine the time to resolution of Long COVID symptoms and the associated factors among the people who were infected with and survived SARS-CoV-2 during the 3rd wave of COVID-19 in Zambia. Information is needed to inform scientific litertaure, policy and development of clinical management guidelines and formulation of health promotion messages for improved long term care of COVID-19 survivors in Zambia.

## Materials and Methods

The methods for this study are described in our previous work [16] as summarised in the sections below.

### Patient and Public Involvement

Participants and the public were not directly involved in the conceptualisation and design of the study. The study design was designed following the call for research proposals on COVID-19 research by the National Science and Technology Council under the strategic research fund. However, selection of study sites was done in consultation with the Ministry of Health and Provincial Health Offices. In addition, selection of study participants was done in consultation with the healthcare facility leaders.

### Study design and setting and participants

The 24-months prospective study was conducted from September 2021 to August, 2023 and employed quantitative (survey) and medical records review as data collection techniques. Corroborating two data collection techniques is likely to increase the validity of the study findings [17,18]. The study was conducted in Lusaka and Southern Provinces of Zambia. A total of seven (7) healthcare facilities were included in the study: 5 from Lusaka province and 2 from Southern province (**Table 1).**

**Table 1:** Study sites.

| **Name of institution** | **Level of care** | **Province** | **n(%)** |
| --- | --- | --- | --- |
| Levy Mwanawasa University teaching hospital | Specialist hospital | Lusaka | 60 (19.8) |
| Kafue General hospital | Regional hospital | Lusaka | 35 (11.6) |
| Chelston hospital | Level 1 hospital | Lusaka | 35 (11.6) |
| Chilenje hospital | Level 1 hospital | Lusaka | 33 (10.8) |
| Chawama hospital | Level 1 hospital | Lusaka | 35 (11.6) |
| Mtendere Mission hospital | Regional hospital | Southern | 50 (16.5) |
| Mazabuka General hospital | Regional Hospital | Southern | 55 (18.1) |
| **Total** |  |  | **303** |

To be included in the study, health facilities must have served as COVID-19 treatment centres during the third wave (June to August 2021). To ensure triangulation of findings, efforts were made to include both urban and rural facilities as well as those providing specialist, second level and primary healthcare. In addition, government/ public and faith-based health facilities were included in the study. Lusaka province has a total of 7 districts, including Lusaka city, the commercial capital and government headquarters, with a total population of 3.19 million [19]; southern province has a total population of 1.85 million [19]. Both provinces provide specialised, second and primary level health care services. Lusaka has 2 specialised, 5 second level and several primary level health care facilities. Southern Province has one specialist hospital, 6 level two, several district hospitals and primary healthcare facilities. In addition, there are private and faith-based healthcare providers in each province. The Ministry of Health (MoH) headquarters and the Zambia National Public Health Institute (ZNPHI)−responsible for the COVID-19 policies, guidelines and disease intelligence−are also located in Lusaka. COVID-19 care services provided in the provinces included screening and testing, contact tracing, laboratory diagnosis, case management and vaccination. Lusaka province was selected because it was the COVID-19 epi-centre due to the huge population and commercial activities. Southern province was selected because it is a transit region for people travelling from the southern region (South Africa, Zimbabwe, Namibia and Botswana)−all of which reported high numbers of COVID cases [20]. Study participants were selected from people who tested positive to and survived COVID-19 during the third wave (June to August, 2021). The third wave was selected based on the large number of people who got infected; it was also chosen for logistical purposes (that is, to make the study feasible). First, a complete list of people who tested positive to COVID-19 during the third wave at each study site was obtained from the line list kept at each health facility. The complete list served as a sampling frame from which potential participants could be sampled. The list contained information about patients including the name, sex, residential and contact details, date of SARS-CoV-2 infection test and diagnosis, condition of patient at the time of diagnosis (symptomatic or not). Next, a stratified random sampling technique was used to select respondents who tested positive to SARS-CoV-2. This approach ensured that both survivors who had mild and/ or no symptoms were selected. To be included into the study, participants needed to:

- Have tested positive to SARS-CoV-2 during the third wave (June-July 2021)
- Be 15 years and above of age
- Have been residing in the selected study area for not less than 3 months
- Have clinical records/ files at the health facility (study site)

Those with active COVID-19 infection, missing data including date of SARS-CoV-2 diagnosis and other sociodemographic and clinical data) in the line list were not eligible to participate. Eligible participants were then contacted on phone and/or email to inform them about the study and confirm their willingness to be recruited in the study. Those who agreed to be included in the study were followed to their homes or any place of their choice for enlisting into the study.

### Variables

The primary outcome variable was the time to resolution of symptoms.

Independent variables or covariates were selected based on priori subject matter knowledge and included:

a. Demographic characteristics: age, sex, number of children, marital status, religious denomination, place of residence
b. Socioeconomic information: level of education, occupation, level of income
c. Medical history: Pre-existing medical conditions, HIV-status, history of admission for acute COVID-19; duration of hospital care
d. Vaccination history: Vaccination status, number of vaccine doses, type of vaccine

### Computation of the time to event variable

To make it feasible to accurately measure the time to resolution of symptoms, several measures were taken. First, key definitions were made including the case definition, date of onset, end date and time to resolution. The case definition of Long COVID was made according to WHO (that is, symptoms appearing 3 or more months post-infection with SARS-CoV-2 infection). The time of onset was defined as the date when the Long COVID symptom was reported or diagnosed. Time of symptom resolution was defined as the date when the participant reported that the symptoms had resolved or when the participant was certified as having no Long COVID symptoms. Right censoring was defined as the symptom that was persistent or remained present at the last follow-up date, or if a participant was lost to follow-up before the symptom resolved. The end date for the participants who were right censored was the end of the follow up period. Time to resolution of symptoms was defined as the duration in days from the date of onset to end date or end of follow up period. To ensure accurate case definition of Long COVID symptoms and computation of the time to resolution of symptoms, data on the date of SARS-CoV-2 infection diagnosis, date of onset of Long COVID symptoms as well as the end date were collected. The Event was defined as the resolution of the symptom or censoring. The Event variable was considered and coded 1 if the symptoms had resolved (coded=1). It was considered 0 if the symptoms had not resolved by the end of the follow up or the participant was lost to follow up (coded=0). The time to resolution of symptoms was computed at the time of data analysis when all the participants had been followed up. Each participant had their own time of onset and end date. This approach ensured that there was no ambiguity in the computation of the time and event variables.

### Data Sources and Procedures

Data were collected by a total of 12 trained and experienced research assistants (2 per research site) under the supervision of 2 field supervisors and the PI. Research assistants were hired from the healthcare facilities where the study was conducted. Hiring and training of data collectors was done by the PI and two field supervisors, after ethical approval, before commencement of the actual data collection process. To be cost-effective, trainings for the research assistants based in Lusaka province were conducted at one central place. Trainings for Kafue and Southern province were conducted separately, at each research site. Each training took a total of five days, in two phases. Phase one comprised theory in research and took 3 days. The main topics covered included: 1) basic principles of research, 2) purpose and objectives of the current study, 3) data collection and interviewing techniques, and 4) informed consent in research involving human subjects, and 5) data collection instruments (survey questionnaire and checklist). Phase two was a field practical and included pre-testing the data collection tools and consent form, and took 2 days. After the field practical, instruments were revised based on the collected and analysed data and feedback from the research assistants. Data were collected at baseline and at a monthly interval up to 18 months. Data were collected from respondents and medical records from the healthcare facilities. Activities at baseline included collection of respondent baseline data, including: a) demographic data, b) date for COVID-19 diagnosis, c) vaccination status, d) date of vaccination, e) previous medical history, e) current symptoms, if any, f) date the symptom was first reported; g) severity of symptoms, h) hospital admission for current symptoms, h) date of admmission; i) duration of admission; j) laboratory and radiological investigations conducted and results,k) medical diagnosis, if known, and l) medical treatment for current symptoms. Activities for subsequent follow-up visits included data collection on: a) respondents’ general well being, b) existing and new symptoms, c) date of onset for new symptoms, d) severity of symptom, e) laboratory/pathology investigations, f) date of symptom resolution, and g) overall outcomes. A paper-based data collection instrument, adapted from the NCL Primary Care COVID-19 recovery questionnaire [22], Derbyshire Post COVID syndrome questionnaire [23], and Post COVID-19 recovery clinic baseline questionnaire [24] were used. To ensure internal validity, the data collection tool was pretested and revised accordingly.

### Potential biases

Possible biases in the study included selection bias, measurement error and confounding due to COVID-19 reinfection. Potential selection bias could have arisen from the non-inclusion of three health facilities that were initially included in the protocol; they were left out due to logistical and administrative challenges. However, data were collected from neighbouring health facilities (already in the protocol) that had similar demographic, socioeconomic and clinic characteristics. Further, a systematic stratified sampling technique was employed to select participants from the line lists maintained in the study sites to ensure that participants with mild and/or no symptoms were recruited. The line list served as a sampling frame from which participants were sampled. In addition, two research team members under the supervision of the PI worked together to select and compile the list of study participants from the sampling frame. To minimise measurement error, a pre-tested data collection instrument was used. Research assistants, trained in data collection techniques, worked in pairs under supervison from the field supervisors. Confounding was controlled for during analysis by adjusting for potential confiunding variables including sex, vaccination status, number of vaccine doses and pre-existing medical conditions, duration of admision and severity of symptoms. Confoudning due to COVID-19 reinfection was difficult to establish during data collection and analysis. However, to ensure uniformity, only participants who tested positive during the third wave of COVID-19 (June, to August, 2021) were included in the sample.

### Study size

We used the choenfeld event-based formula to estimate the sample size. The Cox regression sample-size calculation takes into account the expected hazard ratio, statistical power, the proportion of participants with a characteristic of interest, number of covariates, expected number of events and the duration of follow-up.

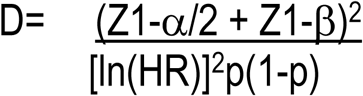

Where:

D= Required number of observed events;

HR = Hazard ratio to be detected;

P= Expected proportion vaccinated;

α= Significance level, usually 0.05;

1-β= Statistical power (80%).

Assuming: HR = 1.5. P= 80%; α= 0.05; 1-β= statistical power (80%); following up period=24 months; 8 covariates, our analysis in RStudio gave us a minimum sample size of 287. Allowing for a 5% loss to follow up=301. We therefore needed a minimum sample of 301 participants.

### Statistical analysis

Data from the survey and medical records was entered into an excel sheet for cleaning up and saving on a password-protected computer. Before the analysis was carried out, the variable for Time to event was created and data calculated and entered in the excel sheet for each participant. Participant follow up time was calculated as: **Time of onset** (=A) to **Time of symptom resolution** (=B). For the participants who were lost to follow up, the last date when they visited the facility or were contacted was recorded as the end date. For those whose symptoms did not resolve by the end of the follow up period, the last date for the follow up period was recorded as the end date. The difference between the Time of symptom resolution and Time of onset (B-A=C) was computed as the follow up time or time to symptom resolution or right censoringuion and recorded in the Time variable for each participant. The Event variable was created and data coded as 0= if the symptoms did not resolve or right censored; and 1= if the symptoms had resolved. Descriptive statistics were computed to summarise sociodemographic and clinical data for all participants. Numerical variables were summarised using the Wilcox test and the median, interquartile range (IQR) and p-value were computed. Categorical variables were summarised using crosstabulations and the percentages and p-values were computed. After descriptive analysis, participants with Long COVID symptoms were identified.

All subsequent analyses were carried on participants who had Long COVID symptoms. The total and median follow up time, number of events, person time, crude incidence rate and its 95% confidence interval (CI) were computed. To compute the incidence rate, we fitted the Poisson regression model using the library(epiR) package (IR <-events / person_time *1000). Next, we fitted the Kaplan-Meier (KM) model to generate the survival curves and visualise their pattern. Survival objects were created using the Surv () function; KM curves were estimated using the survfit() function. After generating the survival curve for the whole sample with Long COVID symptoms, we generated other curves stratified by age, pre-existing medical condition, HIV status and vaccination status. Plots were produced using the survminer package (ggsurvplot) with standardised axes and an abline added to visualise the median time. In addition, the risk table indicating the number of participants with resolved and unresolved symptoms as well as the risk or probability of not having the symptoms resolved at a given point in time, was generated. Next, both univariate and multivariate regression analyses were conducted using the Cox-Proportional Hazards regression model. First, we assessed the univariate association between individual covariates and the outcome variable. Crude Hazard ratios (cHRs), 95% CI and p-values were computed. Next, variables with a p-value of ≤0.2 were then entered into the multivariate Cox-Proportional hazards regression model; adjusted hazard ratios (aHR), 95% CI and p-values were computed. For all the analyses, the outcome variable was dichotomised as 1 = if the symptom had resolved; 0=if the symptom had not resolved, the participant was lost to follow-up or right censored. Model assumptions were assessed by testing the proportional hazards assumptions using the cox.zph () function and Schoenfed residuals; the Chi square goodness of fit test and respective p-values were computed and assessed. P-values > 0.05 were interpreted as no violation of assumptions. The overall model fitness was assessed using the likelihood-ratio test and logrank test; concordance statistic was used to evaluate model discrimination. Akaike’s Information Criterion (AIC) was used to compare competing models. Sensitivity analysis was conducted by comparing the primary final model with two other models: one without the variable on number of vaccine doses and the other one with the age variable categorised into two (below 40 years and 40 years and above). Since we had no missing data, no imputation was done. All analyses were conducted in RStudio version R4.2.1.

### Ethical considerations

Ethical clearance and approval were obtained from the University of Zambia Biomedical Research Ethics Committee (UNZA BREC 2565-2022) (**Supporting information 1**). Permission to conduct the study was granted by the National Health Research Authority. Before participants were recruited into the study, they were contacted on phone for their willngness to participate in the study. Those who accepted were visited by the research assistants and information about the study including the purpose and duration, benefits and risks of participating in the study were explained. Participants were also assured of confidentiality and privacy. Informed consent was obtained from the participants who accepted to particpate in the study. Only participants who signed the consnet form were recruited into the study.

## Results

### Participants

A total of 450 participants were identified as potentially eligible for enrolment into the study. Out of these,330 (73.3%) were contacted for possible enrolment into the study. A total of 303 were enrolled into the study, completed the follow-up and were included in the analysis. Of these, 249 (82.2%) had Long COVID associated symptoms and 154 (17.8%) had no symptoms. A summary of the recruitment algorithm of study participants is shown below (**Fig 1**).

**Figure 1.**
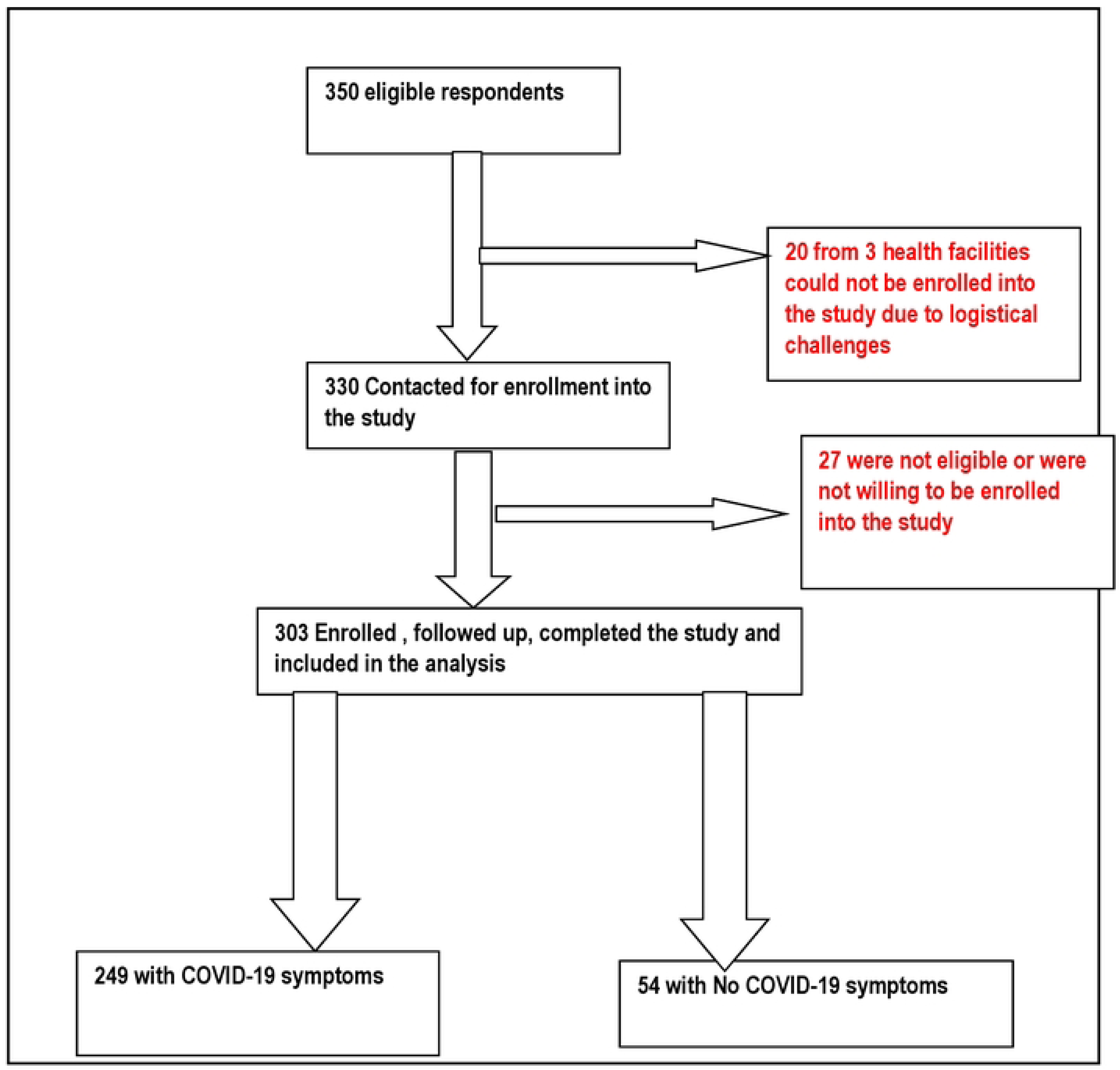
Recruitment algorithm.

### Descriptive statistics

Majority were male (52.8%) with a median age of 35 years (IQR=28-44). Slightly less than half of the sample (47.8) were married and had a median number of children of 2 (IQR=0-3). Majority (61.06%) had attained tertiary education and almost half (48.5%) were in formal employment. Most (55.45%) had an average monthly income of more than ZMW 2000. Regarding religious denomination, almost one third (27.7) of the respondents were Pentecostal. Almost one third (27.06) reported having a pre-existing medical condition. The most prevalent condition was hypertension (41.5%) followed by HIV (28.06%), Diabetes Mellitus (9.76%) and heart disease (6.1%). Majority (62.05%) had received a COVID-19 vaccine: John & Johnson (37.26%), AstraZeneca (22.44%) and Pfizer (2.31%). Almost two-fifths of the respondents (36.6%) reported having received one dose; almost a quarter (23.4%) had received two doses. There was no statistically significant difference between the respondents who had Long COVID and those without with regard to place of residence (p=0.18), age (p=0.31), sex (p=0.11), marital status (p=0.18), number of children (p=0.34), religious affiliation (p=0.95), education level (p=0.87), occupation (p=0.42), income (p=0.31), HIV status (p=0.35), vaccination (p=0.49) and number of vaccine doses (p=0.33). There were more participants with pre-existing medical conditions among those with Long COVID compared with those without Long COVID, although the difference was not statistically significant (p=0.06) **(Table 2).**

**Table 2:**
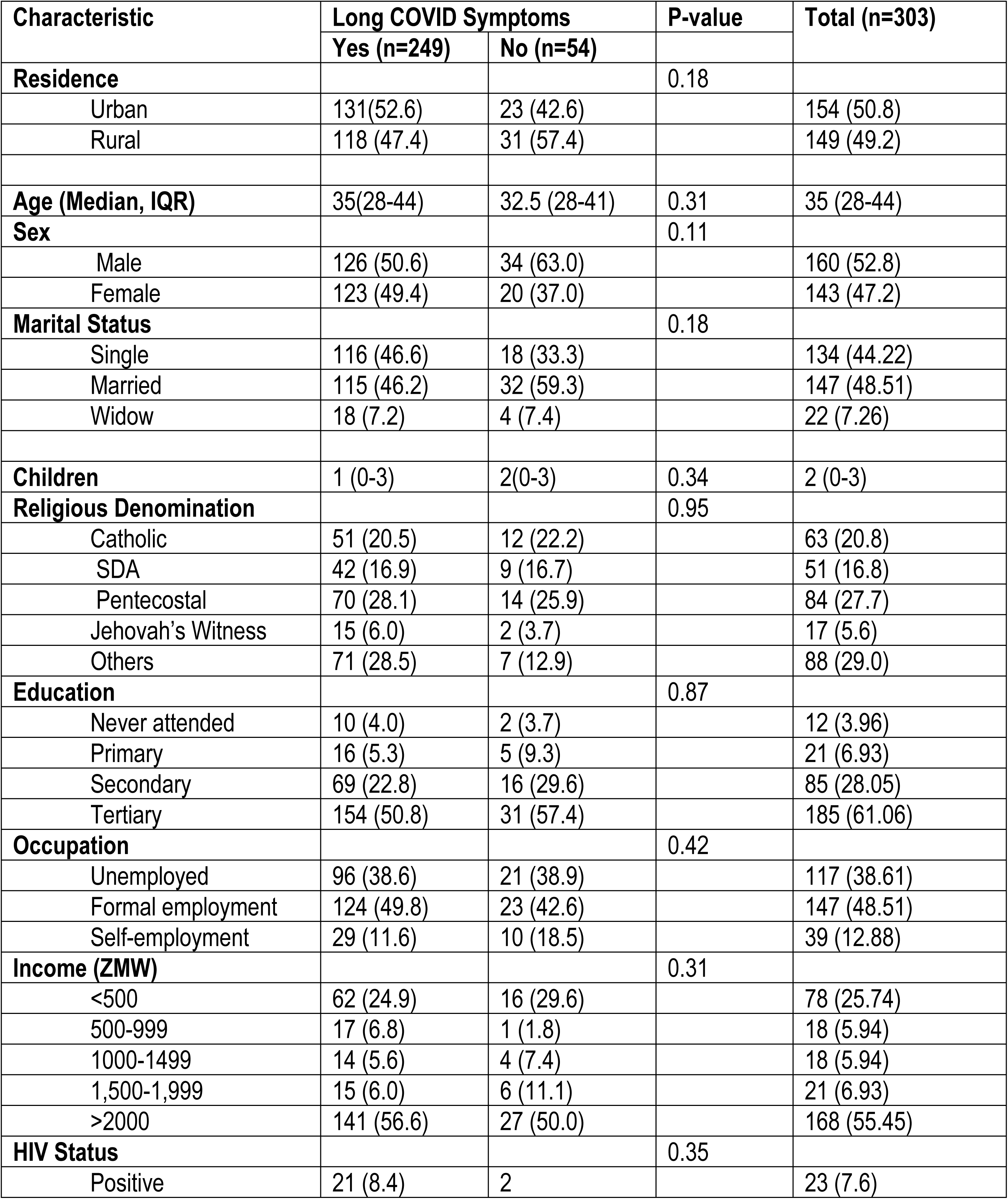
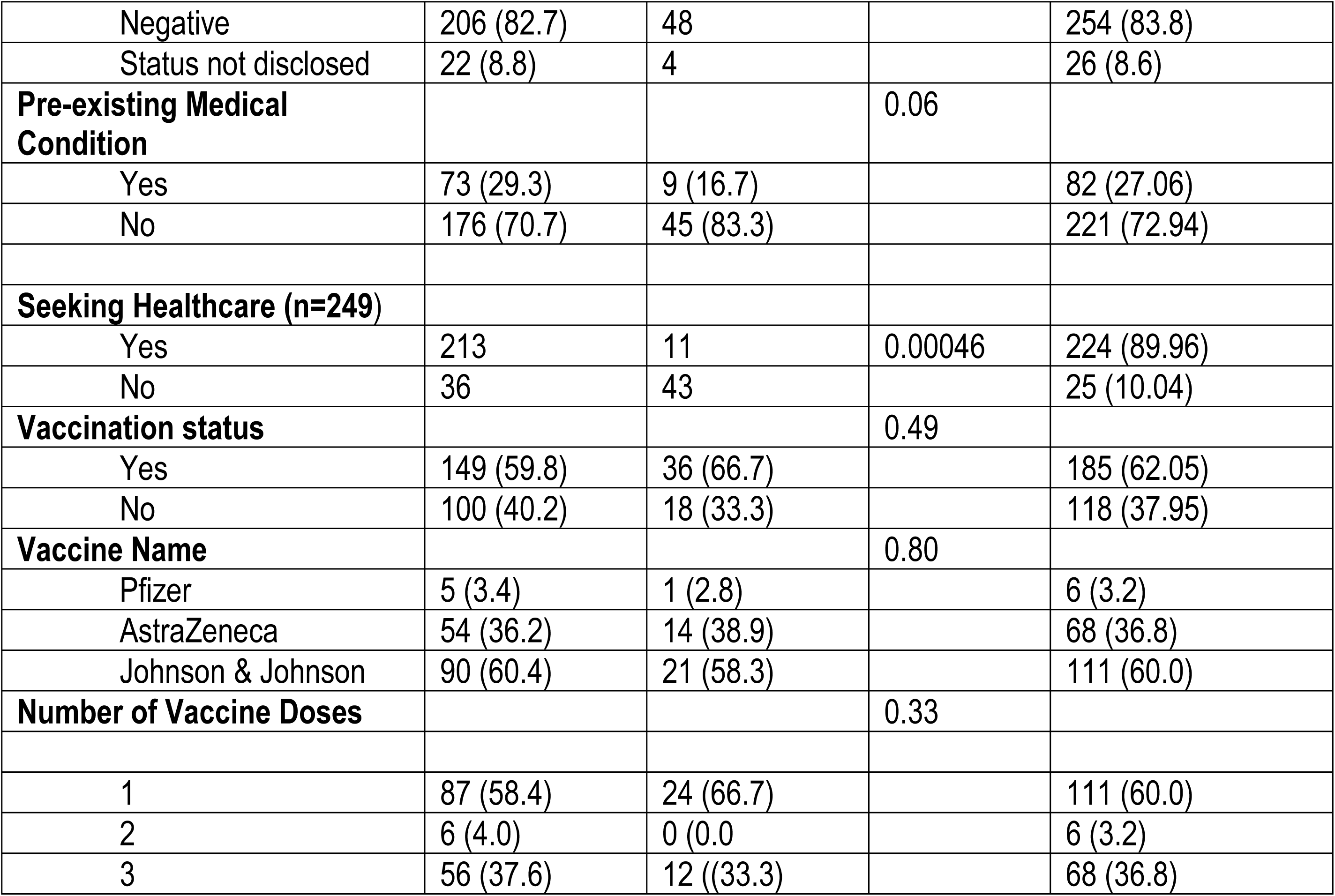
Sociodemographic Characteristics.

| Characteristic | Long COVID Symptoms |  | P-value | Total (n=303) |
| --- | --- | --- | --- | --- |
|  | Yes (n=249) | No (n=54) |  |  |
| **Residence** |  |  | 0.18 |  |
| Urban | 131 (52.6) | 23 (42.6) |  | 154 (50.8) |
| Rural | 118 (47.4) | 31 (57.4) |  | 149 (49.2) |
| **Age (Median, IQR)** | 35 (28-44) | 32.5 (28-41) | 0.31 | 35 (28-44) |
| **Sex** |  |  | 0.11 |  |
| Male | 126 (50.6) | 34 (63.0) |  | 160 (52.8) |
| Female | 123 (49.4) | 20 (37.0) |  | 143 (47.2) |
| **Marital Status** |  |  | 0.18 |  |
| Single | 116 (46.6) | 18 (33.3) |  | 134 (44.22) |
| Married | 115 (46.2) | 32 (59.3) |  | 147 (48.51) |
| Widow | 18 (7.2) | 4 (7.4) |  | 22 (7.26) |
| **Children** | 1 (0-3) | 2 (0-3) | 0.34 | 2 (0-3) |
| **Religious Denomination** |  |  | 0.95 |  |
| Catholic | 51 (20.5) | 12 (22.2) |  | 63 (20.8) |
| SDA | 42 (16.9) | 9 (16.7) |  | 51 (16.8) |
| Pentecostal | 70 (28.1) | 14 (25.9) |  | 84 (27.7) |
| Jehovah's Witness | 15 (6.0) | 2 (3.7) |  | 17 (5.6) |
| Others | 71 (28.5) | 7 (12.9) |  | 88 (29.0) |
| **Education** |  |  | 0.87 |  |
| Never attended | 10 (4.0) | 2 (3.7) |  | 12 (3.96) |
| Primary | 16 (5.3) | 5 (9.3) |  | 21 (6.93) |
| Secondary | 69 (22.8) | 16 (29.6) |  | 85 (28.05) |
| Tertiary | 154 (50.8) | 31 (57.4) |  | 185 (61.06) |
| **Occupation** |  |  | 0.42 |  |
| Unemployed | 96 (38.6) | 21 (38.9) |  | 117 (38.61) |
| Formal employment | 124 (49.8) | 23 (42.6) |  | 147 (48.51) |
| Self-employment | 29 (11.6) | 10 (18.5) |  | 39 (12.88) |
| **Income (ZMW)** |  |  | 0.31 |  |
| <500 | 62 (24.9) | 16 (29.6) |  | 78 (25.74) |
| 500-999 | 17 (6.8) | 1 (1.8) |  | 18 (5.94) |
| 1000-1499 | 14 (5.6) | 4 (7.4) |  | 18 (5.94) |
| 1,500-1,999 | 15 (6.0) | 6 (11.1) |  | 21 (6.93) |
| >2000 | 141 (56.6) | 27 (50.0) |  | 168 (55.45) |
| **HIV Status** |  |  | 0.35 |  |
| Positive | 21 (8.4) | 2 |  | 23 (7.6) |
| Negative | 206 (82.7) | 48 |  | 254 (83.8) |
| Status not disclosed | 22 (8.8) | 4 |  | 26 (8.6) |
| **Pre-existing Medical Condition** |  |  | 0.06 |  |
| Yes | 73 (29.3) | 9 (16.7) |  | 82 (27.06) |
| No | 176 (70.7) | 45 (83.3) |  | 221 (72.94) |
| **Seeking Healthcare (n=249)** |  |  |  |  |
| Yes | 213 | 11 | 0.00046 | 224 (89.96) |
| No | 36 | 43 |  | 25 (10.04) |
| **Vaccination status** |  |  | 0.49 |  |
| Yes | 149 (59.8) | 36 (66.7) |  | 185 (62.05) |
| No | 100 (40.2) | 18 (33.3) |  | 118 (37.95) |
| **Vaccine Name** |  |  | 0.80 |  |
| Pfizer | 5 (3.4) | 1 (2.8) |  | 6 (3.2) |
| AstraZeneca | 54 (36.2) | 14 (38.9) |  | 68 (36.8) |
| Johnson & Johnson | 90 (60.4) | 21 (58.3) |  | 111 (60.0) |
| **Number of Vaccine Doses** |  |  | 0.33 |  |
| 1 | 87 (58.4) | 24 (66.7) |  | 111 (60.0) |
| 2 | 6 (4.0) | 0 (0.0) |  | 6 (3.2) |
| 3 | 56 (37.6) | 12 ((33.3) |  | 68 (36.8) |

### Long COVID Symptoms

A total of 249 (82.2%) respondents had Long COVID-related symptoms. The most common symptoms were respiratory and cardiac symptoms (61.04%) followed by digestive symptoms (38.96%), general body symptoms including fatigue and general body weakness, neurological and reproductive symptoms. Table 3 below shows the type of Long COVID symptoms and the number of participants who experienced them **(Table 3).**

**Table 3:**
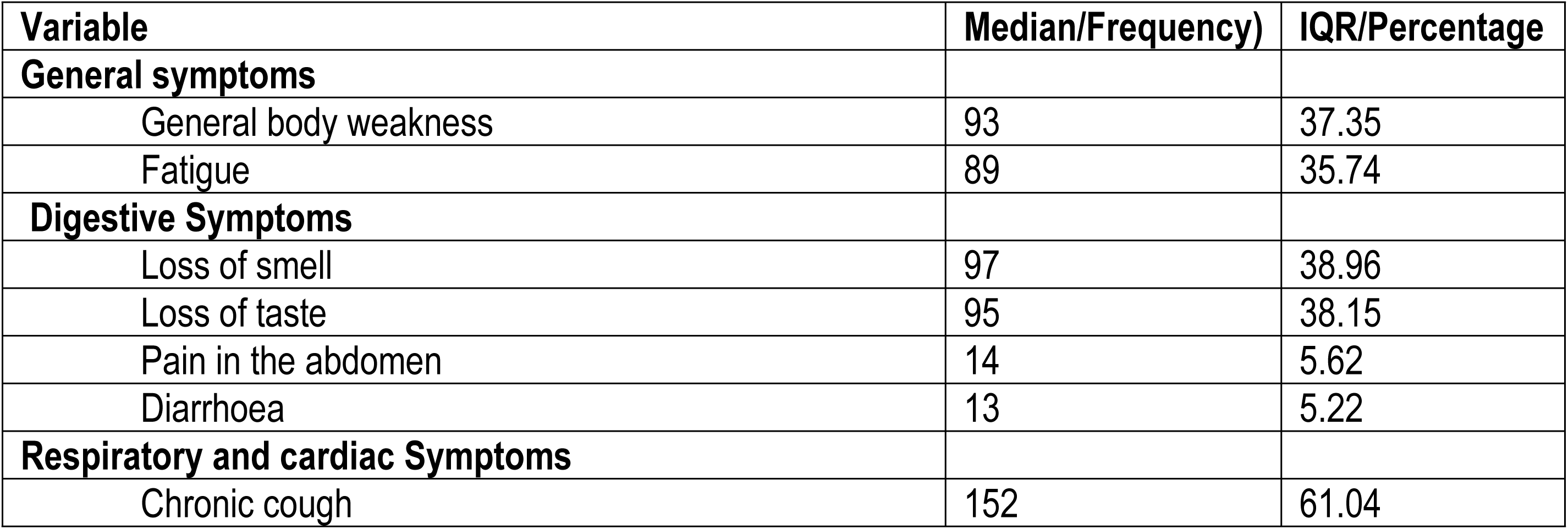
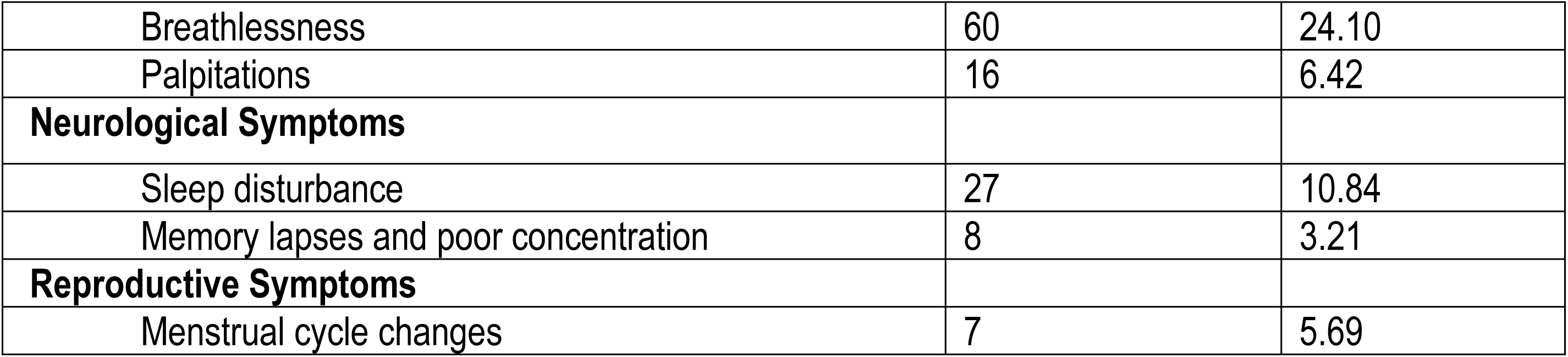
Long COVID symptoms.

### Number of events and incidence rate

A total of 236 events occurred during the follow-up period over 1, 798 person-months, giving a crude incidence rate of 131.3 events per 1000 person-months (95%CI: 115.2 - 148.7) or 1, 575.6 events per 1000 person years (95%CI: 1,363.2 - 1,784.4).

### Survival analysis using Kaplan Meier Model

Fig 2 shows the Kaplan–Meier (KM) curve to estimate the time from diagnosis to resolution of symptoms among the respondents who had Long COVID symptoms. The crude median time from diagnosis of Long COVID symptoms to resolution was 6 (IQR: 5-8) months. Comparisons on survival time were made on age (Fig 3, p-value=0.07); pre-existing medical conditions (Fig 4, p-value=0.07) and vaccination status (Fig 5, p-value=0.046)

**Figure 2.**
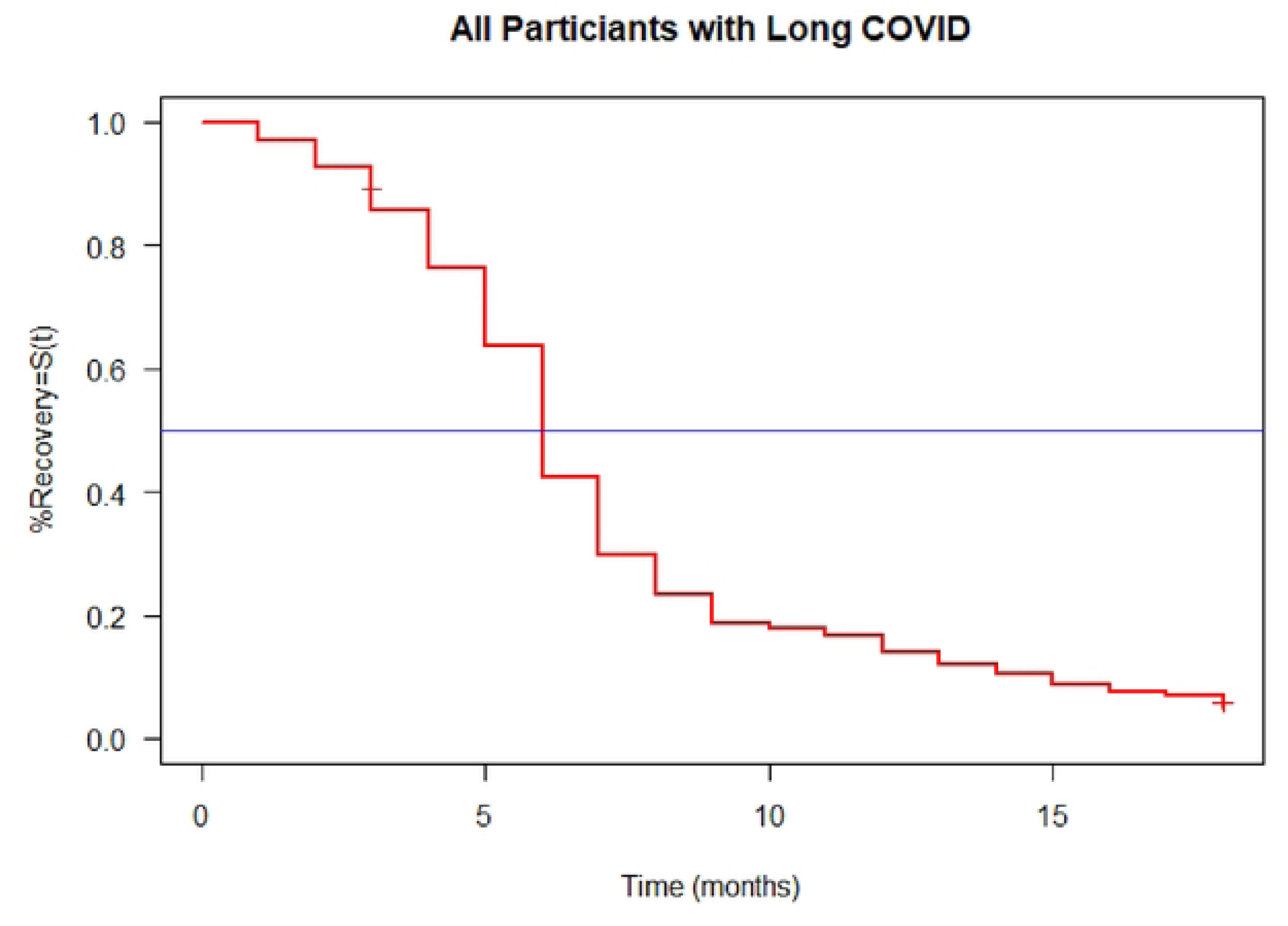

**Fig 3.**
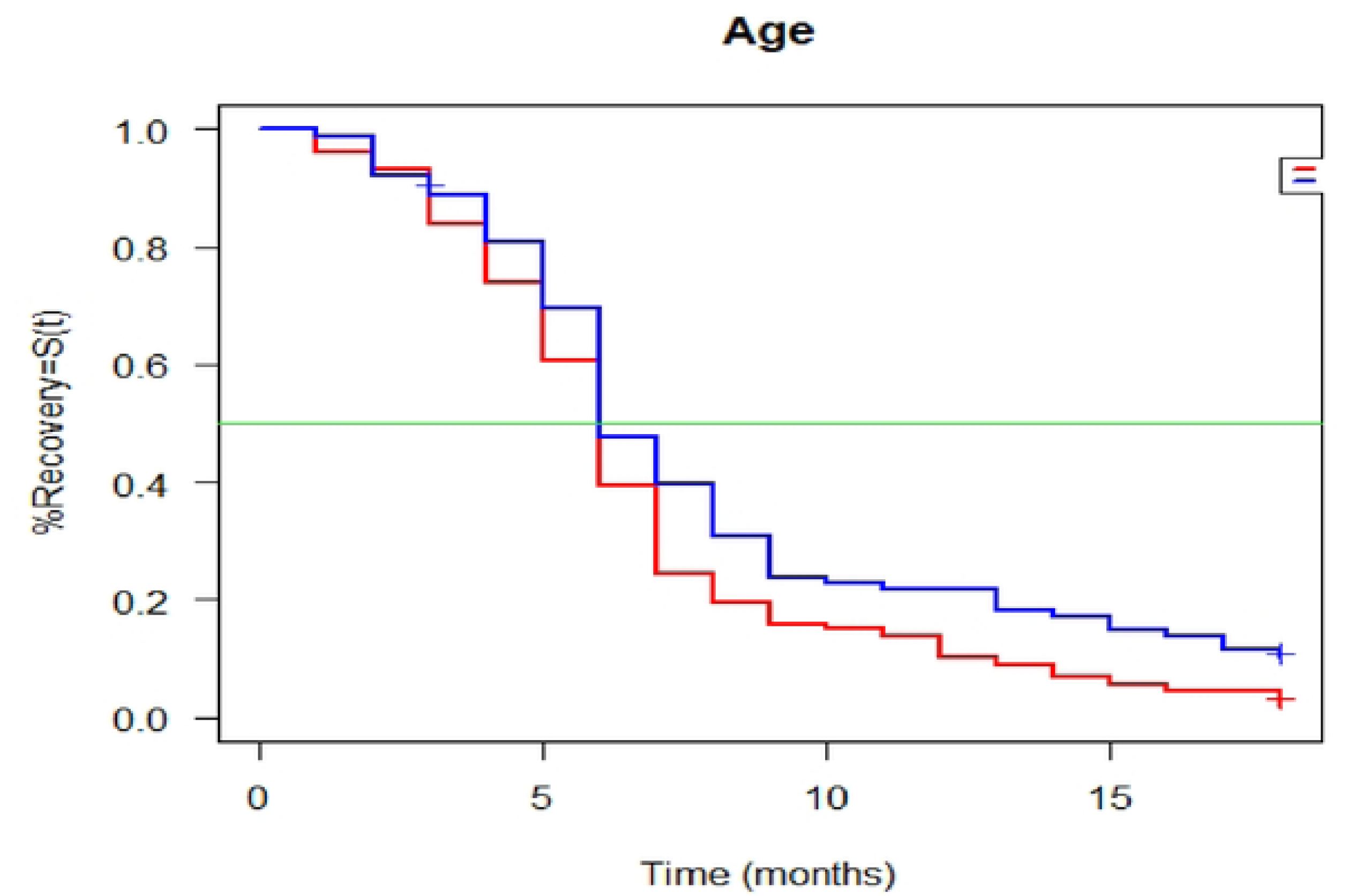

**Fig 4:**
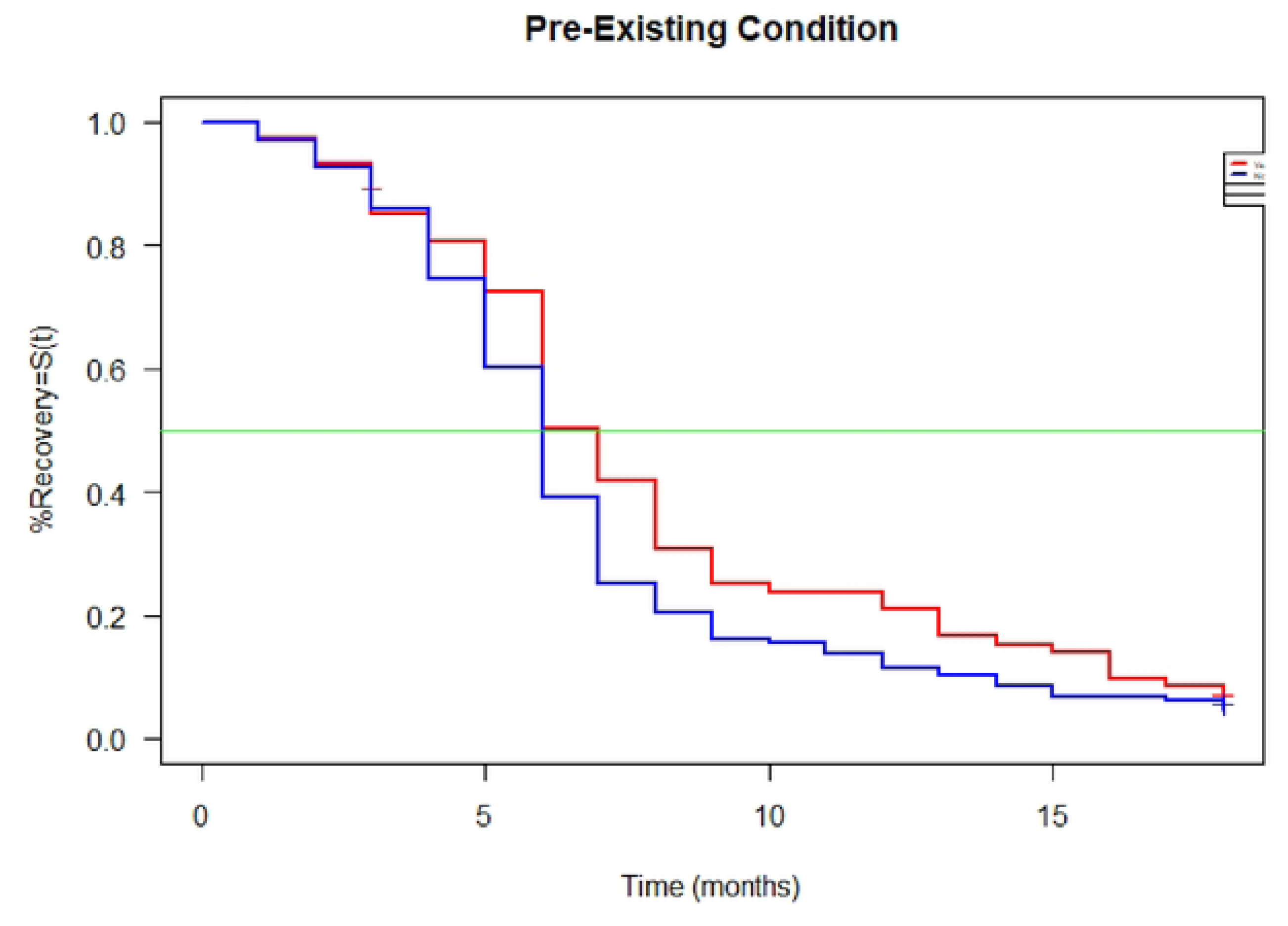

**Fig 5.**
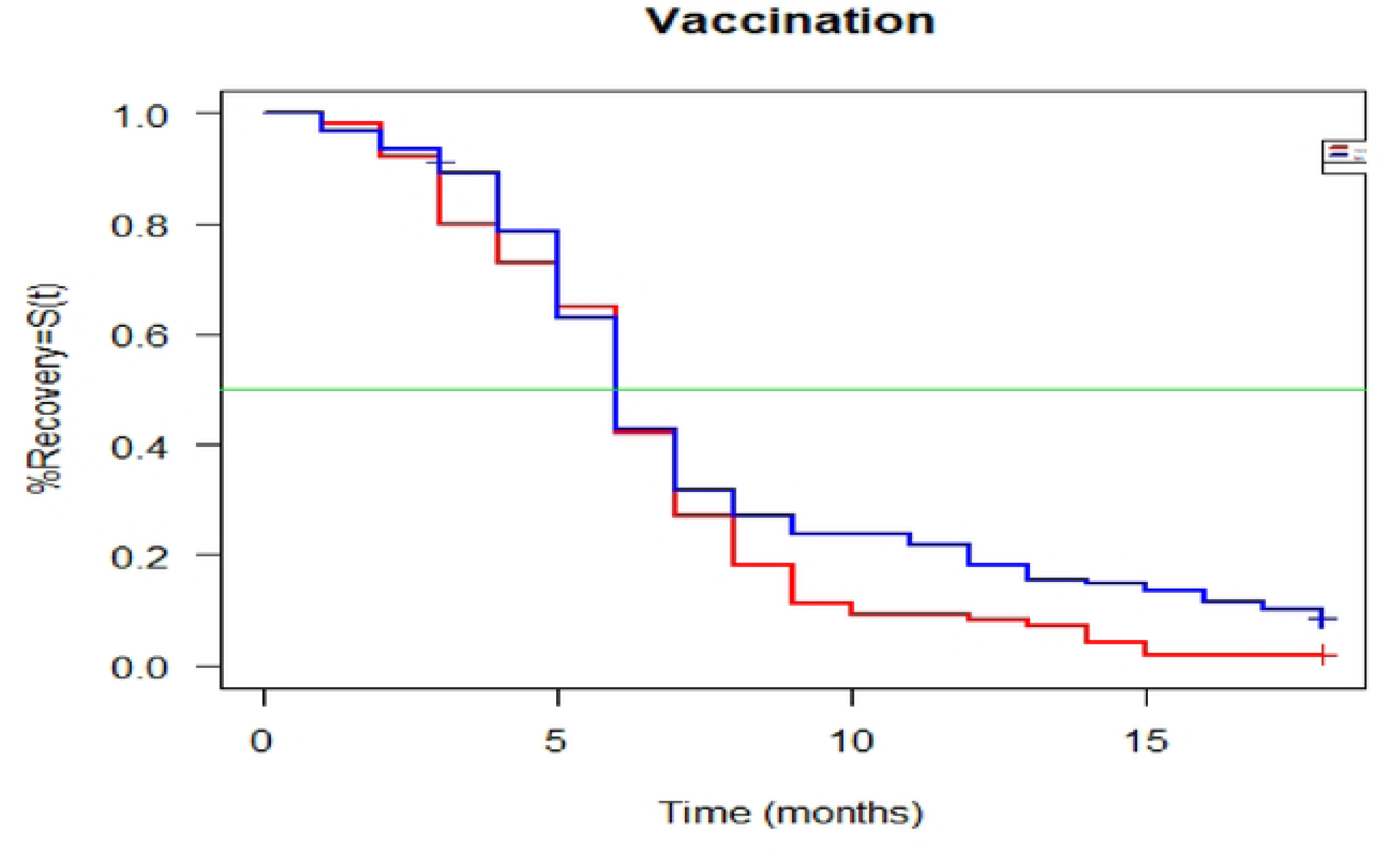

### Symptom resolution analysis

The table below shows the number of participants with resolved and unresolved symptoms as well as the risk or probability of not having the symptoms resolved at a given point in time. There was a total of 249 respondents with Long COVID symptoms at baseline. A total of 7 recoveries occurred at month 1, giving a probability of having unresolved symptoms of 97.2% (95%CI [95.2% to 99.2%]). At month 2, a total of 242 respondents still had symptoms and 11 events occurred, resulting in a probability of having unresolved symptoms of 92.8% (95% [89.6% to 96.0%]). Going into month 3, a total of 231 respondents still had symptoms; 18 recovered and 1 case was censored, resulting in a probability of unresolved symptoms of 85.5% (95%CI [81.3% to 90.05]). At month 18, a total of 17 respondents still had unresolved symptoms; 5 recovered, resulting in a probability of unresolved symptoms of 4.8% (95%CI [28.0% to 84.0%]). A total of 13 participants had unresolved symptoms at the end of the follow up period and were right censored **(Table 4)**.

**Table 4:** Symptom Resolution Analysis.

| Time | Number at risk | Events | Probability | 95%CI |  |
| --- | --- | --- | --- | --- | --- |
| 1 | 249 | 7 | 0.972 | 0.952 | 0.993 |
| 2 | 242 | 11 | 0.928 | 0.896 | 0.960 |
| 3 | 231 | 18 | 0.855 | 0.813 | 0.900 |
| 4 | 212 | 23 | 0.763 | 0.712 | 0.817 |
| 5 | 189 | 31 | 0.637 | 0.580 | 0.700 |
| 6 | 158 | 53 | 0.424 | 0.366 | 0.489 |
| 7 | 105 | 31 | 0.299 | 0.247 | 0.361 |
| 8 | 74 | 16 | 0.234 | 0.187 | 0.293 |
| 9 | 58 | 12 | 0.186 | 0.143 | 0.241 |
| 10 | 46 | 2 | 0.178 | 0.136 | 0.232 |
| 11 | 44 | 3 | 0.165 | 0.125 | 0.219 |
| 12 | 41 | 6 | 0.141 | 0.104 | 0.192 |
| 13 | 35 | 5 | 0.121 | 0.087 | 0.169 |
| 14 | 30 | 4 | 0.105 | 0.073 | 0.151 |
| 15 | 26 | 4 | 0.089 | 0.059 | 0.132 |
| 16 | 22 | 3 | 0.0076 | 0.049 | 0.118 |
| 17 | 19 | 2 | 0.069 | 0.043 | 0.108 |
| 18 | 17 | 5 | 0.048 | 0.028 | 0.084 |

### Univariate Cox-proportional hazards regression analysis

To identify the individiual factors associated with the time to resoltution of Long COVID symptom, we fitted the Cox-Proportional hazards regression model. First, we examined the univariate association of the selected covariates with the outcome variable and computed the crude hazard ratios (cHRs) and their corresponding p-values. There was a significant crude association between and age and resolution of symptoms (cHR= 0.988, 95% CI: 0.978 - 0.998, p-value=0.02), number of children (cHR=0.932, 95% CI: 0.877 - 0.990, p-value=0.024) and vaccination status (cHR=1.302, 95%CI: 1.00 - 1.693, p=0.046). The association between the outcome and pre-existing medical condition (cHR=1.294, 95% CI: 0.975 - 1.717, p-value=0.07) did not reach the level of significance. Other variables showed no significant association with the time to resolution of symptoms (p-value >0.05) **(Tables 5).**

**Table 5:**
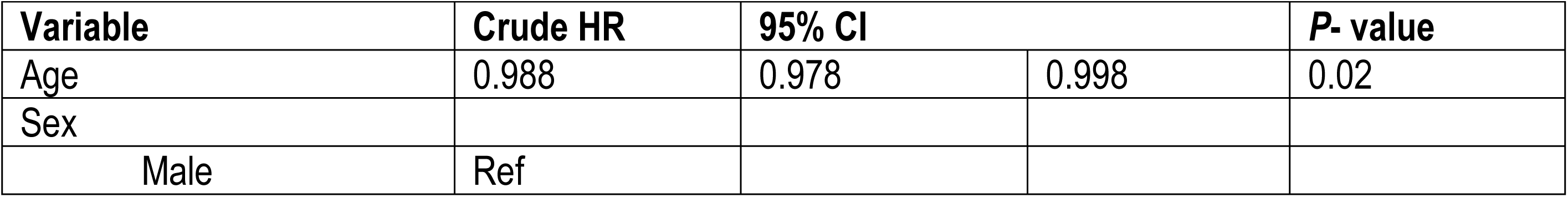
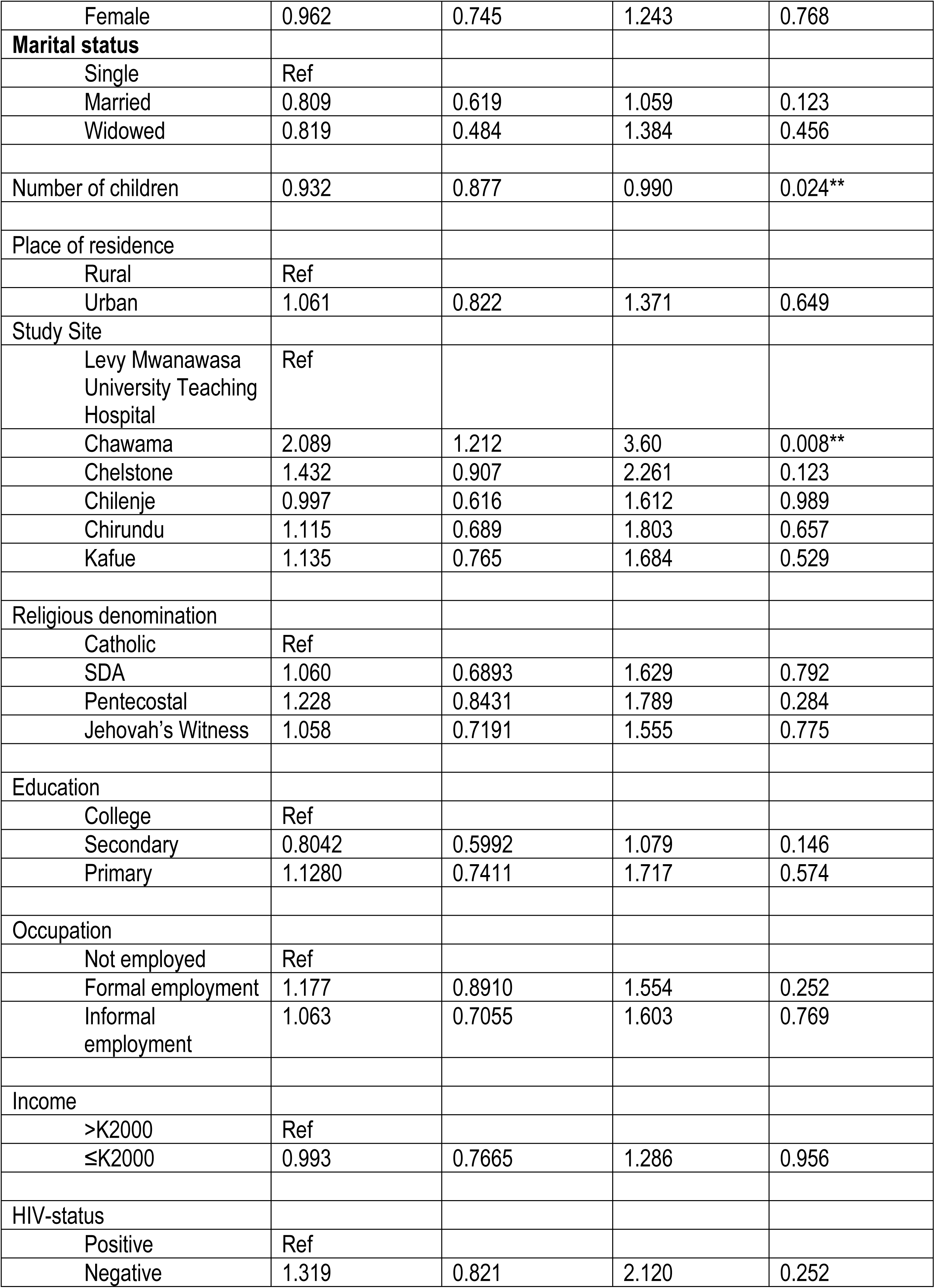
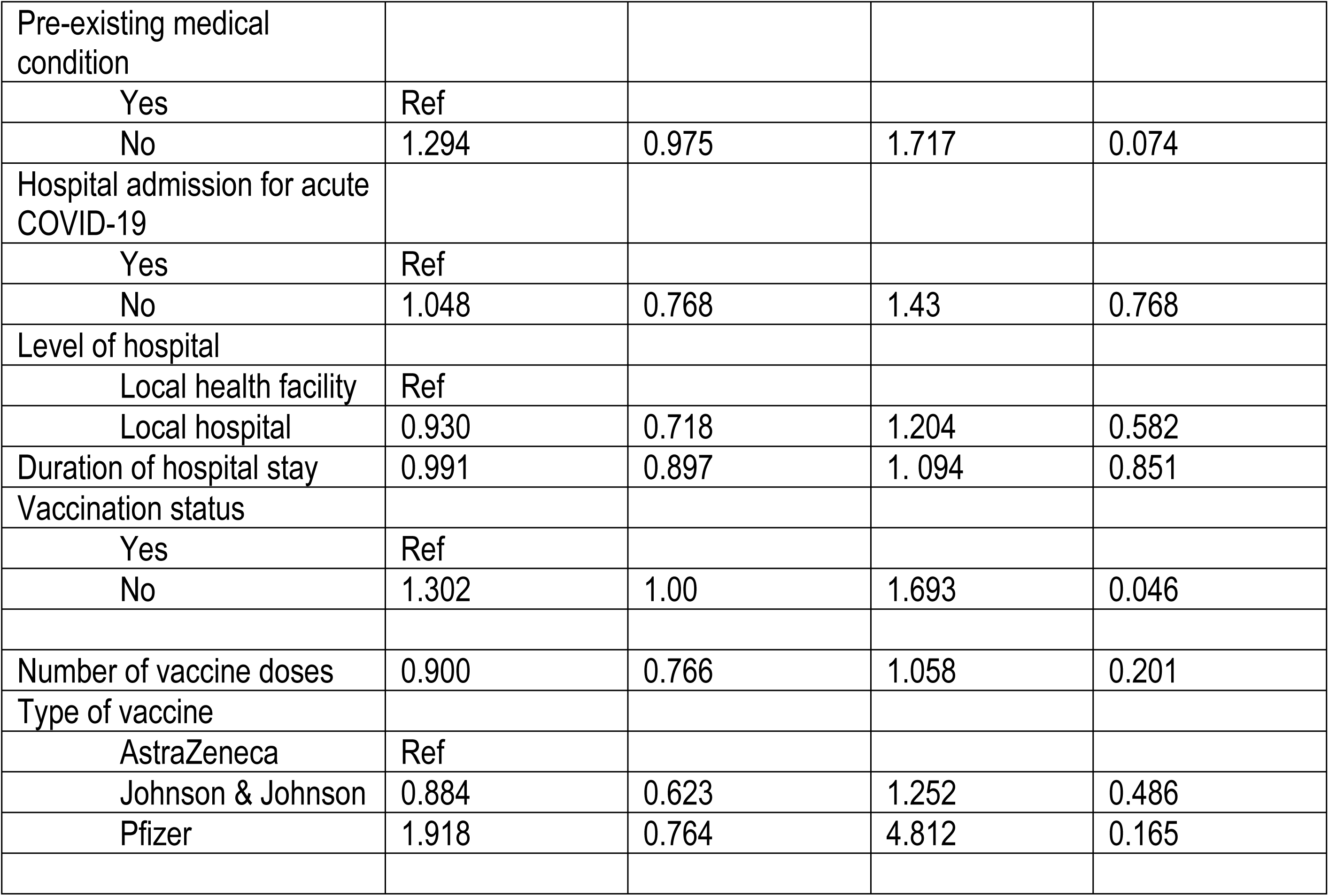
Univariate Cox-proportional hazards regression analysis.

| Variable | Crude HR | 95% CI |  | P- value |
| --- | --- | --- | --- | --- |
| Age | 0.988 | 0.978 | 0.998 | 0.02 |
| Sex |  |  |  |  |
| Male | Ref |  |  |  |
| Female | 0.962 | 0.745 | 1.243 | 0.768 |
| **Marital status** |  |  |  |  |
| Single | Ref |  |  |  |
| Married | 0.809 | 0.619 | 1.059 | 0.123 |
| Widowed | 0.819 | 0.484 | 1.384 | 0.456 |
| Number of children | 0.932 | 0.877 | 0.990 | 0.024** |
| Place of residence |  |  |  |  |
| Rural | Ref |  |  |  |
| Urban | 1.061 | 0.822 | 1.371 | 0.649 |
| Study Site |  |  |  |  |
| Levy Mwanawasa University Teaching Hospital | Ref |  |  |  |
| Chawama | 2.089 | 1.212 | 3.60 | 0.008** |
| Chelstone | 1.432 | 0.907 | 2.261 | 0.123 |
| Chilenje | 0.997 | 0.616 | 1.612 | 0.989 |
| Chirundu | 1.115 | 0.689 | 1.803 | 0.657 |
| Kafue | 1.135 | 0.765 | 1.684 | 0.529 |
| Religious denomination |  |  |  |  |
| Catholic | Ref |  |  |  |
| SDA | 1.060 | 0.6893 | 1.629 | 0.792 |
| Pentecostal | 1.228 | 0.8431 | 1.789 | 0.284 |
| Jehovah's Witness | 1.058 | 0.7191 | 1.555 | 0.775 |
| Education |  |  |  |  |
| College | Ref |  |  |  |
| Secondary | 0.8042 | 0.5992 | 1.079 | 0.146 |
| Primary | 1.1280 | 0.7411 | 1.717 | 0.574 |
| Occupation |  |  |  |  |
| Not employed | Ref |  |  |  |
| Formal employment | 1.177 | 0.8910 | 1.554 | 0.252 |
| Informal employment | 1.063 | 0.7055 | 1.603 | 0.769 |
| Income |  |  |  |  |
| >K2000 | Ref |  |  |  |
| ≤K2000 | 0.993 | 0.7665 | 1.286 | 0.956 |
| HIV-status |  |  |  |  |
| Positive | Ref |  |  |  |
| Negative | 1.319 | 0.821 | 2.120 | 0.252 |
| Pre-existing medical condition |  |  |  |  |
| Yes | Ref |  |  |  |
| No | 1.294 | 0.975 | 1.717 | 0.074 |
| Hospital admission for acute COVID-19 |  |  |  |  |
| Yes | Ref |  |  |  |
| No | 1.048 | 0.768 | 1.43 | 0.768 |
| Level of hospital |  |  |  |  |
| Local health facility | Ref |  |  |  |
| Local hospital | 0.930 | 0.718 | 1.204 | 0.582 |
| Duration of hospital stay | 0.991 | 0.897 | 1.094 | 0.851 |
| Vaccination status |  |  |  |  |
| Yes | Ref |  |  |  |
| No | 1.302 | 1.00 | 1.693 | 0.046 |
| Number of vaccine doses | 0.900 | 0.766 | 1.058 | 0.201 |
| Type of vaccine |  |  |  |  |
| AstraZeneca | Ref |  |  |  |
| Johnson & Johnson | 0.884 | 0.623 | 1.252 | 0.486 |
| Pfizer | 1.918 | 0.764 | 4.812 | 0.165 |

### Multivariate Cox-proportional hazards regression analysis

To assess the multivariate association between the covariates and the outcome variable, we fitted the multivariate Cox-proportional hazards regression model. Variables that showed association with the outcome variable with a p-value of value ≤0.2 (that is, age, number of children, previous medical condition, vaccination status and number of vaccine doses) were loaded into the model for analysis. In addition, hospital admission was included in the model despite having a p-value of 0.768. The decision was made based on priori knowledge that admission in a hospital is an indicator of the severity of a medical condition and has been shown to be associated with the time to symptom resolution. Adjusted hazard ratios (aHRs), 95% CI and their corresponding p-values are shown in Table 6 below (**Table 6).**

**Table 6:** Multivariate Cox-Proportional Hazard regression analysis.

| Variable | Adjusted HR | 95% CI |  | P- value |
| --- | --- | --- | --- | --- |
| Vaccination |  |  |  |  |
| No |  |  |  |  |
| Yes | 1.71 | 1.02 | 2.86 | 0.04** |
| Age | 0.99 | 0.98 | 1.01 | 0.46 |
| Children | 0.96 | 0.89 | 1.03 | 0.22 |
| Occupation |  |  |  |  |
| Unemployed | Ref |  |  |  |
| Formal employment | 1.13 | 0.849 | 1.51 | 0.40 |
| Informal employment | 0.97 | 0.64 | 1.50 | 0.92 |
| Pre-existing medical condition |  |  |  |  |
| Yes |  |  |  |  |
| No | 1.23 | 0.90 | 1.69 | 0.19 |
| Number of vaccine doses | 1.19 | 0.88 | 1.62 | 0.25 |
| Hospital admission for acute COVID care | 0.88 | 0.62 | 1.24 | 0.47 |
| No |  |  |  |  |
| Yes | 1.13 | 0.81 | 1.59 | 0.47 |

Table 6 shows the Multivariable Cox-proportional hazards regression results. Vaccination status was significantly associated with the time to resolution of the Long COVID symptoms (aHR= 1.71, 95% CI: 1.02 - 2.86, p=0.04) after adjusting for age, number of children, occupation, pre-existing medical conditions, number of vaccine doses and hospital admission during the acute COVID-19 phase. Participants who were vaccinated had a 71% higher rate (hazard) of symptom resolution compared to the unvaccinated particpants. Other variables did not show statistically significant association (p>0.05) with the time to resolution of symptoms. The model parameters (likelihood ratio test (LRT) = 14.3, p = 0.05, Wald test=13.8 on 8 df, p = 0.05) and log-rank test =13.92 on 8 df, p = 0.05)) show that the overall model was statistically significant. The concordance of 0.57 (se = 0.023) indicates a modest discriminatory ability.

### Model assumptions

Table below shows the Chi square and p-values for each variable included in the model assumption analysis. The p-values greater than 0.05 (p>0.05) confirm that there was no statistical evidence of violation of the proportional hazards assumptions for any of the variables included in the model (**Table 7**).

**Table 7:** Model assumptions.

| Variable | Chi- square | p-value | Interpretation |
| --- | --- | --- | --- |
| Age | 0.409 | 0.523 | No evidence of violation |
| Children | 0.056 | 0.813 |  |
| Occupation | 1.136 | 0.567 | No evidence of violation |
| Pre-existing Medical Condition | 0.554 | 0.457 | No evidence of violation |
| Vaccination | 3.984 | 0.061 | No significant violation |
| Number of Vaccine Doses | 1.141 | 0.285 | No evidence of violation |
| Hospital Admission | 1.448 | 0.257 | No evidence of violation |

### Sensitivity analysis

Table 8 below shows the results of our sensitivity analysis and confirms the robustness of the findings. Vaccination remained significantly associated with faster resolution of Long COVID symptoms across the three sensitivity analyses. The aHR was 1.71 in the primary model, it dropped to 1.32 after excluding the covariate on number of vaccine doses and returned to 1.69 when the age variable was categorised. Although the strength of association was reduced when the covariate on the number of vaccine doses was removed from the model, the direction of association remained unaltered and the association remained statistically significant. Categorisation of age produced an estimate similar to that of the primary model, supporting the robustness of the findings to alternative specifications of the age (Table 8).

**Table 8:** Sensitivity analysis.

| Model | aHR for Vaccination | 95% CI |  | P- value |
| --- | --- | --- | --- | --- |
| Primary model | 1.71 | 1.02 | 2.86 | 0.04 |
| Without Vaccine doses | 1.32 | 1.006 | 1.74 | 0.04 |
| With age categorized | 1.69 | 1.01 | 2.82 | 0.05 |

## Discussion

This study aimed to determine the time to resolution of Long COVID symptoms and the associated factors among the people who were infected with and survived SARS-CoV-2 during the 3rd wave of COVID-19 in Zambia. The findings showed that, although most respondents experienced a range of Long COVID symptoms including respiratory and cardiac symptoms, digestive symptoms, general body symptoms, neurological and sexual and reproductive symptoms including chronic cough, palpitations, chest pains and breathless, general body weakness, fatigue and muscle pains, loss of smell and taste, sleep disturbance, memory lapses and poor concentration as well as menstrual cycle changes among female respondents, most participants had recovered. The median time to resolution of symptoms was 6 months. Vaccination status was significantly associated with the time to resolution of symptoms, after adjusting for age, number of children, occupation, pre-existing medical conditions, number of vaccine doses and hospital admission during the acute COVID-19 phase. At any given time during the follow up period, vaccinated participants had a 71% higher rate of symptom resolution compared to the unvaccinated particpants. Other variables did not show statistically significant association with the time to resolution of symptoms.

These findings corroborate previous studies which reported Long COVID and unresolved symptoms post the acute phase among the COVID-19 survivors. Our study shows that the most common symptoms associated with Long COVID affected the respiratory and cardiac, digestive, musculoskeletal, neurological, and sexual and reproductive systems. The most common symptoms included chronic cough, palpitations, chest pains and breathlessness, general body weakness, fatigue and muscle pains, loss of smell and taste, sleep disturbance, memory lapses and poor concentration as well as menstrual cycle changes among female respondents. Although most symptoms were mild, several respondents reported changes in their functional status. For example, body weakness, joint pains and fatigue made it difficult for them to carry out their daily routines such as walking, bathing, washing and other self-care activities. Difficulties in breathing, chronic cough, chest pains, palpitations and other associated respiratory and cardiac symptoms affected the functional status of the survivors. These findings are consistent with those reported in previous studies. For examples, a systematic review and meta-analysis on the long-term sequelae of COVID-19 infection conducted by Rahmati and colleagues [25] reported persistence of unresolved symptoms including dyspnoea, fatigue, insomnia, loss of smell, loss of taste and anxiety. The findings are also consistent with those reported in a study conducted by Malambo and colleagues [26] on clinical characteristics and factors associated with Long COVID among post-acute COVID-19 clinic patients in Zambia. These authors reported presence of long-term clinical effects of COVID-19 on survivors and that these clinical effects affected functional status of the survivors. These findings have implications for future research, healthcare policy, and patient care ─including continued clinical monitoring of the survivors in order to assess and attend to their health needs, even after they have been discharged from health facilities

Although most respondents reported having unresolved Long COVID symptoms, majority recovered during the follow up period. Only 13 (5.2%) respondents did not have their symptoms resolved by the end of the follow-up period. The median time to resolution of symptoms was 6 months. The survival curve and risk table clearly show that the proportion of participants with unresolved symptoms as well as well as the probability of having unresolved symptoms continued reducing over time. This finding is consistent with a study conducted in Switzerland by Ballouz and colleagues [27] which reported a steady reduction in the proportion of infected individuals with unresolved symptoms. These authors reported that 22.9% (95% CI: 20.4% to 25.6%) of the study participants did not fully recover by six months. They also showed that the proportion of individuals who reported unresolved symptoms decreased from 18.5% (95%CI: 16.2% to 21.1%) at 12 months to 17.2% (95%CI: 14.0% to 20.8%) at 24 months after infection. This finding is consistent with the World Health Organisation (WHO) which shows that symptoms of post COVID-19 condition generally improve over time, typically 4-9 months [28]. WHO also indicates that approximately 15% of the people still have symptoms at 12 months [29].

Consistent with previous studies that have reported the protective effect of vaccination on COVID-19 outcomes, our findings showed a significant association between vaccination status and probability of recovery. Participants who were vaccinated had a 71% higher rate of symptom resolution compared to the unvaccinated particpants after adjusting for age, number of children, occupation, pre-existing medical conditions, number of vaccine doses and hospital admission during the acute COVID-19 phase. Sensitivity analysis showed that the association between vaccination and time to resolution was sensitive to the number of vaccine doses. However, the number of doses did not diminish the association or change the direction of the relationship. This finding is important as it clearly shows the importance of not only reciving the vaccine but also reciving the recommended number of doses. This finding is consistent with those reported by Guimarães and colleagues [30] in their study on vaccination and COVID-19: impact on Long-COVID which showed that vvaccination against SARS-CoV-2 significantly reduces its incidence and symptom burden. The finding is also consistent with various studies that have demonstrated lower frequencies of fatigue, cough, dyspnea, anxiety, depression, memory dysfunction, and brain fog among vaccinated individuals compared with unvaccinated peers, with effects persisting for up to 18 months [31–33].

Our study showed that, in the univariate Cox-proportional hazards regression analysis, age and number of children were each statistically associated with the rate of recovery from Long COVID symptoms without adjustment for other covariates. At any given time during the follow up period, there was a 1.2% reduction in the rate of recovery with a unit increase in the participant’s age in years. There was 6.8% reduction in the rate of recovery with an increase in the number of children. However, the association disappeared after adjusting for other covariates in the multivariate model. The reason for this finding is not clear; the finding could suggest that the observed unadjusted association may be partly explained by differences in other participant characteristics. Further research is needed to investigate this relationship. Nevertheless, this finding is important as it collaborates other studies that have reported the association between age and the time to resolution of Long COVID symptoms [34]. The finding is also consistent with the Global Burden of Disease Long COVID [35] which showed that older adults have an increased risk of having unresolved symptoms. Similarly, a systematic review with meta-analysis on the isolated effect of age on the risk of COVID-19 severe outcomes by Romero and others [30] showed that the risk of in-hospital mortality increased with age by 5.7% (effect size (ES) in-hospital mortality=1.057, 95% CI: 1.038 to 1.054). The risk of case fatality increased with age by 7.4% (ES case fatality=1.074, 95% CI: 1.061 to 1.087). The study also reported that the risk of hospitalisation increased by 3.4% per age year (ES=1.034, 95% CI: 1.021 to 1.048).

Consistent with previous studies that reported a significant association between pre-existing medical conditions and the probability of recovery [ 36,37], our findings showed that a significant association between pre-existing medical conditions (comorbidities) and the time to resolution of symptoms before we adjusted for other covariates in the model. Respondents with comorbidities had a 29.4% higher rate of recovery compared with those that had comorbidities. Our findings show that 27.1% of the respondents with Long COVID had pre-existing medical conditions; the most prevalent ones were hypertension followed by HIV, Diabetes Mellitus and heart disease. However, this relationship was not significant at multivariate level. The reason for the insignificant multivariate association is not clear. Further research is required to investigate this relationship. Possible explanations for this finding may be that the significant association at univariate level was due to confounding. The univariate association may have resulted from the influence of other variables associated with both comorbidities and time to recovery. The other reason could be that comorbidities may be correlated with other covariates such as age. Several studies have demonstrated the relationship between age and comorbidities; as people advance in age, the probability of developing comorbidities such as diabetes mellitus, hypertensive heart disease, renal disease and others increases. For example, in their multicohort analyses of 108, 810 adults on the role of age in the association of chronic conditions and multimorbidity with incident activity of daily living (ADL) reported that chronic conditions were positively associated with age [38].

Possible limitations of the study should be acknowledged. Although we collected data on comorbidities including HIV status, we did not collect data on mental health (anxiety, depression) and lifestyle factors. We recommend future studies to investigate the relationship between mental health and the time to resolution of symptoms. Moreover, although our findings clearly show a statistically significant association between vaccination status and the time to resolution of Long COVID symptoms, the causal relationship can not be inferred due to the observational study design. Studies with an experimental design are needed to establish the causal relationship. Finally, since the study was conducted in selected health facilities in Zambia, its findings may have limited generalizability outside Zambia due to differences in geographical, epidemiological clinical characteristics.

Despite these potential limitations of the study, we belive these findings provide important insight into the time to resolution of symptoms and associated factors among the SARS-CoV-2 survivors in Zambia. Our study methods and analytical approach are replicable. The model parameters (likelihood ratio test (LRT), log-rank test and concordance clearly show that the overall model was robust enough to predict the time to resolution of symptoms with modest discriminatory ability to distinquish participants with the event and those without. Further, our analysis clearly shows that the model assumptions were not violated.. Finally, our sensitivity analysis further supports the robustness of the findings. These tests confirm the validity of our study findings.

## Conclusion

This study aimed to determine the time to resolution of Long COVID symptoms and the associated factors among the people who were infected with and survived SARS-CoV-2 during the 3rd wave of COVID-19 in Zambia. Although most respondents experienced a range of unresolved symptoms, including respiratory and cardiac symptoms, digestive symptoms, general body symptoms, neurological and sexual and reproductive symptoms, majority recovered. The median time to resolution of symptoms was 6 months. The study found that vaccination status was statistically associated with the time to resolution of symptoms after adjusting for other covariates age, number of children, occupation, comorbidities, number of vaccine doses and hospital admission during the acute COVID-19 phase. Participants who were vaccinated had higher (71%) hazards of symptom resolution compared to the unvaccinated particpants. This finding suggests that vaccination was associated with a shorter time to resolution of Long COVID symptoms and highlight the importance of vaccination status in understanding recovery from Long COVID. However, causation can not be ascertained due to the observational design of our study. Studies with an experimental design are needed to establish the causal relationship.

## Data Availability

Data is available and will be shared on reasonable request

## Contributors

CS led the design of the study, drafting of the study protocol and implementation of the study, led the analysis and drafted this manuscript. EK, PIS and NM contributed to the development of the data collection instruments, enlisting the study sample, and coordinated data collection process. VM contributed to the revision of the manuscript. All authors read and approved the final draft of the manuscript.

## Disclaimer

The funder had no influence on the design and implementation of the study, nor on the draft of this manuscript.

## Patient and public involvement

Patients and/or the public were not involved in the design, or conduct, or reporting, or dissemination plans of this research. Refer to the Methods section for further details.

## Patient consent for publication

Ethical clearance and approval were obtained from the University of Zambia Biomedical Research Ethics Committee (UNZA BREC 2565-2022)

